# Effectiveness of Cardiopulmonary Bypass Priming with Albumin, Gelofusine and Crystalloids with Retrograde Autologous Priming in Preventing Microcirculatory Dysfunction in Patients during Coronary Artery Bypass Graft Surgery - (PRIME) a Double-Blind Randomized Trial

**DOI:** 10.1101/2025.04.01.25325060

**Authors:** Anne M. Beukers, Jord Seegers, Nikki van Haasteren, Meike Brouwers, Ruben J. Bosch, Anita M. Tuip-de Boer, Charissa E. van den Brom, Susanne Eberl, Gudrun Kunst, David M.P. van Meenen, Carolien S.E. Bulte, Stephan A. Loer, Alexander B.A. Vonk, PRIME study group, MICROnet consortium

**Affiliations:** Department of Anesthesiology, Amsterdam University Medical Centre, Amsterdam, the Netherlands; Department of Intensive Care, Tergooi Hospital, Hilversum, the Netherlands; University of Amsterdam, Amsterdam, the Netherlands; Vrije Universiteit, Amsterdam, the Netherlands; Department of Clinical Perfusion, St. Antonius Hospital, Nieuwegein, the Netherlands; Laboratory for Experimental Intensive Care and Anesthesiology (LEICA), Amsterdam University Medical Centre, Amsterdam, the Netherlands; Department of Intensive Care Medicine, Amsterdam University Medical Centre, Amsterdam, the Netherlands; Department of Anesthesiology, King’s College Hospital, London, the United Kingdom; Department of Cardiothoracic Surgery, Amsterdam University Medical Centre, Amsterdam, the Netherlands

**Keywords:** Microcirculation, Colloid, Crystalloid, Glycocalyx, Colloid oncotic pressure, Endothelium

## Abstract

**Introduction:** Cardiopulmonary bypass (CPB) can cause endothelial dysfunction, leading to edema formation, disturbed microcirculatory perfusion, and impaired tissue oxygenation. Together, these factors contribute to organ dysfunction in patients undergoing cardiac surgery. We hypothesis that the composition of CPB prime fluids influences endothelial function and prevents microcirculatory dysfunction. We assessed the impact of three different CPB prime fluid strategies on microcirculatory perfusion in adult patients undergoing coronary artery bypass graft surgery.

**Methods:** Thirty-four patients were subjected to CPB primed with 1500 mL of either albumin/ringers, gelofusine/ringers, or solely ringers plus retrograde autologous priming (RAP). All three priming solutions included 100 mL of mannitol. The primary outcome was perfused vessel density (PVD) assessed by sublingual microcirculatory imaging at 5 time points (after anesthesia induction, aortic cross-clamping, weaning from CPB, upon intensive care unit arrival (ICU), and 24 hrs after ICU arrival) as main marker of microcirculatory perfusion. Secondary outcomes included total vessel density, proportion of perfused vessels, microvascular flow-, and heterogeneity index (as additional markers of microcirculatory perfusion), colloid oncotic pressure (COP), albumin, glycocalyx shedding-, endothelial-, inflammation- and renal injury markers, hemoglobin, hematocrit, hemolysis index, transfusion volumes, fluid balance and fluid requirements.

**Results:** CPB immediately impaired PVD across all groups (albumin/ringers estimated mean difference between baseline and arrival ICU -7.56 95% CI [-11.53 to -3.59] mm mm^−2^, gelofusine/ringers -4.10 [-7.53 to -0.67] mm mm^−2^, and ringers plus RAP -3.77 [-6.64 to -0.90] mm mm^−2^), persisting until ICU arrival without differences between groups. In patients receiving gelofusine/ringers COP was preserved after aortic cross clamping, and was associated with lower intraoperative fluid requirements and fluid balances compared to those receiving albumin/ringers or ringers plus RAP. Levels of inflammatory (interleukin-6) and endothelial damage markers (angiopoietin-2) were higher in patients receiving albumin/ringers compared with those receiving gelofusine/ringers, and ringers plus RAP.

**Conclusion:** All of the three CPB priming strategies - albumin/ringers, gelofusine/ringers or ringers plus RAP – similarly induced perioperatieve microcirculatory dysfunction in patients undergoing CABG surgery. These findings suggest that none of these prime fluid strategies confers protection for microcirculatory perfusion during cardiac surgery. Albumin priming resulted in increased inflammatory markers at 24 hrs after surgery.

**Trial registration:** ClinicalTrials.gov, NCT05647057. Registered on 04/25/2023. ClinicalTrials.gov <u>PRS: Record Summary NCT05647057</u>, all items can be found in the protocol.

## Introduction

Patients following cardiac surgery with cardiopulmonary bypass (CPB) face increased risk for organ dysfunction, particularly renal and pulmonary injury, which is associated with increased morbidity and mortality.^1–5^ The microcirculatory perfusion, relying on its convective (e.g., flow) and diffusive (e.g., distance from red blood cell to cells) capacities and essential in preserving organ function, decreases by 20% from the onset of CPB, and might contribute to the development of organ dysfunction following on-pump cardiac surgery.^6–11^

Microcirculatory dysfunction during cardiac surgery is linked to a decrease in capillary density and perfusion, resulting in a reduced ability of the microcirculation to deliver oxygen to tissues.^12,13^ The underlying causes of a decreased perfused capillaries and capillary density are increased endothelial permeability and vascular leakage, both consequences of systemic inflammation, endothelial activation, hemodilution, and hemolysis during CPB.^1,7,10,14–16^ Microcirculatory perfusion during CPB could be influenced by prime fluid strategy, which is known as major contributor to hemodilution during CPB. Therefore, selecting an optimal prime fluid strategy, that minimizes hemodilution, preventing vascular leakage and preserving microcirculatory function, may be crucial. Studies have shown that using albumin as CPB priming preserves colloid oncotic pressure (COP), reduces interstitial fluid accumulation, and lowers the risk of myocardial injury compared to crystalloids.^17–19^ In rats on CPB, albumin led to less pulmonary edema, renal injury, inflammation and glycocalyx degradation compared to hydroxyethyl starch as priming.^20^ The impact of gelofusine, a more affordable synthetic colloid, on microcirculatory perfusion remains unexamined. Additionally, strategies designed to reduce net priming volume - and consequently hemodilution - such as retrograde autologous priming (RAP), have been directly associated with reduced transfusions.^21^ However, there is no optimal strategy for CPB priming. The PRIME study aimed to compare the effects of albumin/ringers, gelofusine/ringers, or ringers combined with RAP as CPB prime fluid strategies on microcirculatory perfusion in patients undergoing on-pump coronary artery bypass graft surgery (CABG). In this study, we hypothesized that either albumin/ringers or gelofusine/ringers would better preserve microcirculatory perfusion compared to ringers plus RAP as CPB priming.

## Methods

### Study design

The PRIME study was a single-center, double-blind randomized clinical trial to compare three different prime fluid strategies in patients receiving CABG surgery with CPB at the Amsterdam University Medical Center, The Netherlands. We previously published the protocol and statistical analysis plan.^22^ The institutional review board approved the trial (approval number 23-0036). Written informed consent was obtained from all patients. The CONSORT guidelines were followed in drafting of this manuscript (EQUATOR network).^23^

### Patients

The trial enrolled adult patients receiving CABG surgery with CPB. Exclusion criteria were emergency surgery; aortic surgery; valve surgery; combined procedures valve and CABG surgery; known allergy for human albumin or gelofusine; the use of crystalloid cardioplegia.

### Randomization

Patients were randomized in a 1:1:1 ratio to a CPB priming strategy of albumin/ringers, gelofusine/ringers, or solely ringers with RAP. The local investigators randomized patients using Castor (EDC, 2020). The randomization sequence was generated using random block sizes of three, six and nine. Patients were randomized the day before surgery to allow sufficient time for preparation of CPB priming. Patients were replaced if the procedure or date of surgery changed, that led to the use of different cardioplegia. The study was designed as a physiological proof-of-principle study to evaluate the effect of CPB priming on microcirculatory perfusion, without the potential interference of other types of cardioplegia. Further details regarding the randomization process are provided in Supplementary Material, and Table S1. Inclusions continued until sample size per arm was achieved. The patients and investigators were blinded for the intervention. Attending anesthesiologist, cardiothoracic surgeon and perfusionist were aware of the perfusion strategy in individual patients, but they were not aware of the primary outcome.

### Interventions

Patients were randomized into three priming strategies: 1500 mL of gelofusine/ringers (750 mL modified fluid gelatin (Braun Melsungen AG, Germany), 650 mL Ringer’s solution (Baxter BV, Utrecht, Netherlands) and 100 mL mannitol (15%, Baxter BV, Utrecht, Netherlands)); 1500 mL of albumin/ringers (200 mL human albumin (20%, Sanquin, Amsterdam, Netherlands), 1200 mL Ringer’s solution (Baxter BV, Utrecht, Netherlands) and 100 mL mannitol (15%, Baxter BV, Utrecht, Netherlands)), or 1500 mL of Ringers plus RAP (1400 mL Ringer’s solution (Baxter BV, Utrecht, Netherlands) and 100 mL mannitol (15%, Baxter BV, Utrecht, Netherlands) with RAP. RAP was applied to a maximum volume of 475 mL in case of a body surface area (BSA) > 1.7 m^2^ and 375 mL in case of BSA < 1.7 m^2^ provided that systolic blood pressure will remain > 90 mmHg. If additional fluids were needed during CPB to maintain optimal perfusion, the displaced prime fluid as result of RAP was used prior to the fluid protocol.

In case of hemodilution, defned as a hematocrit below 0.24 L L^−1^, RBC transfusions were used. In case of a cardiac index <2.2 L min^−1^ m^−2^, patients received noradrenaline 0.02–0.20 mcg kg^−1^ min^−1^, with phenylephrine boluses of 50–500 mcg as bridge to noradrenaline. Before and after CPB, patients received Plasmalyte based on the decision of the anesthesiologist. Besides the intervention groups, no additional colloid fuids were given in the perioperative process.

### Outcomes

The primary outcome was the change in perfused vessel density (PVD), reflecting microcirculatory diffusion capacity, collected using a non-invasive sidestream dark field (SDF) video microscopy (USB3 with SDF technology, Microvision Medical, Amsterdam, the Netherlands) for sublingual microcirculatory measurements. Videos of 4 seconds were recorded from 3 different sublingual sites per timepoint. The acquired images were double stored offline for Microcirculation Imaging Quality Scoring analyses, and manually analyzed with AVA 3.2.^24^ The technique is performed according to the international guidelines for sublingual microcirculatory measurements, and is detailed in the study protocol previously published.^22,25^ The primary outcome was measured at five consecutive time points: after anesthesia induction, after aortic cross clamping, after weaning from CPB, after arrival on the intensive care unit and 24 hours after ICU arrival.

Secondary outcomes included total vessel density (TVD), proportion of perfused vessels (PPV), heterogeneity index, microvascular flow index (MFI), COP, albumin, glycocalyx shedding-, endothelial-, inflammation- and renal injury markers: syndecan-1, heparan sulphate, thrombomodulin, angiopoietin-2 (Ang-2), interleukin-6 (IL-6), tumor necrosis factor alpha (TNF-α), neutrophil gelatinase-associated lipocalin (NGAL), hemoglobin, hematocrit, hemolysis index, perioperative use of transfusion products, fluid balance and fluid requirements. The data collection process and measurement of secondary outcomes are outlined in our previously published study protocol.^22^

Perioperative blood samples were collected from an arterial line into dipotassium ethylenediaminetetraacetic acid (K_2_EDTA), heparin, and citrate containing tubes. A total of 80 mL of blood was drawn per patient for analysis. Plasma was stored at -80°C for further testing. COP was measured using an Osmomat 050 (Gonotec, Berlin, Germany), with plasma injected into an oncotic cell. Osmotic pressure differences were detected via electronic pressure sensors and displayed in mmHg. Glycocalyx shedding-, endothelial-, infammatory-, and renal injury markers are measured using ELISA (SEA565hu and SEB966hu, Cloud-clone Corporation, Hubei, China) and a Luminex platform (Biotechne) in accordance with the manufacturer’s instructions and corrected for corresponding hematocrit levels.

### Anesthesia and CPB protocol

Anesthesia was induced using intravenous sufentanil, propofol, lidocaine, midazolam, and ketamine combined with rocuronium bromide and maintained by continuous propofol infusion (4–6 mg kg^−1^ h^−1^) and continuous sufentanil infusion (0.5–1 mcg kg^−1^ h^−1^). The lungs were ventilated with a tidal volume of 6–8 ml kg^−1^. An O2-air mixture with an FiO_2_ of 0.4–0.5 and a positive end-expiratory pressure of 5 cm H_2_O was be used. All patients received cefazolin (2 g), dexamethasone (0.5 mg kg^−1^), and clemastine (2 mg). All patients received tranexamic acid before initiation of CPB (10 mg kg^−1^) followed by a continuous infusion of 1–2 mg kg^−1^ h^−1^. Preoperative volume losses were replaced using Plasmalyte. A C5 or a S5 heart–lung machine (LivaNova Nederland NV, Amsterdam, Netherlands) with a centrifugal pump and a heater-cooler device, HCU 40 (Getinge Nederland, Hilversum, Netherlands), was used for CPB. Te bypass circuit consisted of a phosphorylcholine-coated tubing system (LivaNova Nederland BV, Amsterdam, Netherlands) with an oxygenator (Inspire 8F) and arterial line, a soft shell venous reservoir (BMR 1900, Medtronic/LivaNova Nederland NV, Amsterdam, Netherlands), and a HVR cardiotomy reservoir (Medtronic/LivaNova Nederland NV, Amsterdam, Netherlands). Te extracorporeal circuit was primed based on the study groups. A 24-French arterial cannula was placed in the ascending aorta and a venous cannula (36 French/46 French) was placed in the right atrium after administration of heparin (300 IE kg^−1^). Venting of the aorta was achieved by an aortic root cannula. When the activated clotting time exceeded 400 s (Hemochron Signature Elite, Edison, USA), it was considered safe to initiate bypass. Blood fow during mild hypothermic (34 °C) CPB was kept between 2.2 and 2.6 L min^−1^ m^−2^. Myocardial protection was achieved by autologous warm blood cardioplegia, and weaning from CPB was started when the rectal temperature had reached 36 °C. A cell- saving device (Xtra™ Autotransfusion System, LivaNova Nederland BV, Amsterdam, Netherlands) was used for retransfusion of pericardial shed blood. After weaning from CPB, administration of protamine reverses heparin in a 0.6:1 fashion and was monitored with HepCon and ROTEM analysis.

### Sample size

Sample size calculation was based on the decrease in PVD from induction of anesthesia to CPB initiation of 5 mm mm^-2^, with a standard deviation (SD) of 4 mm mm^-2^.^9,26,27^ A sample size of 24 patients (8 patients per group) would have 80% statistical power to show a maximal decrease in PVD of 5 mm mm^-2^ (standard deviation 4), using an alpha of 0.05, over three groups with 5 consequtive measurements. Thirty patients were enrolled (10 in each group) to allow for an anticipated dropout of 20%. Secondary study measurements were explorative, hence no sample size calculation was needed.

### Statisitical analysis

For the primary analysis, the change in PVD over time was compared between groups using linear mixed effects models. Models included time as fixed effect and patient was included as random effect. If a difference was found, means for different time points were compared with pairwise comparisons with Bonferroni-adjusted significance levels (p value multiplied by the number of test in the pairwise comparisons, and using an alpha of 0.05). Results of the pairwise comparisons of the means for different time points were rather explorative, since the primary outcome is set on the overall between-group difference.

Data were expressed as percentages (%), as mean ± standard deviation or median [interquartile range] in case of non-normally distributed variables. Normality was checked by means of normal-probability plots (boxplot, Q-Q plot) and the Kolmogorov Smirnov and Shaprio-Wilk tests. The change in TVD, PPV, heterogeneity index, MFI, COP, albumin, syndecan-1, heparan sulphate, thrombomodulin, Ang-2, IL-6, TNF-α, NGAL, hemoglobin, hematocrit, and hemolysis index over time was compared between groups using linear mixed effect models. If a difference was found, means for different time points were compared with pariwise comparions, as described for the primary outcome. For comparison of normally distributed continuous variables, means were compared using one-way ANOVA. For comparison of continuous non-normally distributed variables medians were compared using the Kruskal-Wallis test. Categorical variables were compared by chi-square or Fisher’s exact test as appropriate. R version 4.3.2 (R, Vienna, Austria) was used for all statistical analyses.

## Results

### Patients

Patients were recruited from July 10^th^ 2023, to July 8^th^ 2024. 62 patients were eligible for study participation. In total 48 patients were randomized, 13 patients were replaced due to changes in surgical procedure or date of surgery that led to the use of different cardioplegia. 1 patients was excluded from the final analysis due to inadequate microcirculation imaging quality scoring. 34 patients were included in the final analysis. (Figure 1 and Table S3) More information on randomization can be found in the Supplementary Materials. (Figure 1) Baseline patients characteristics, preoperative laboratory values, and surgical characteristics were comparable between the study groups. (Table 1)

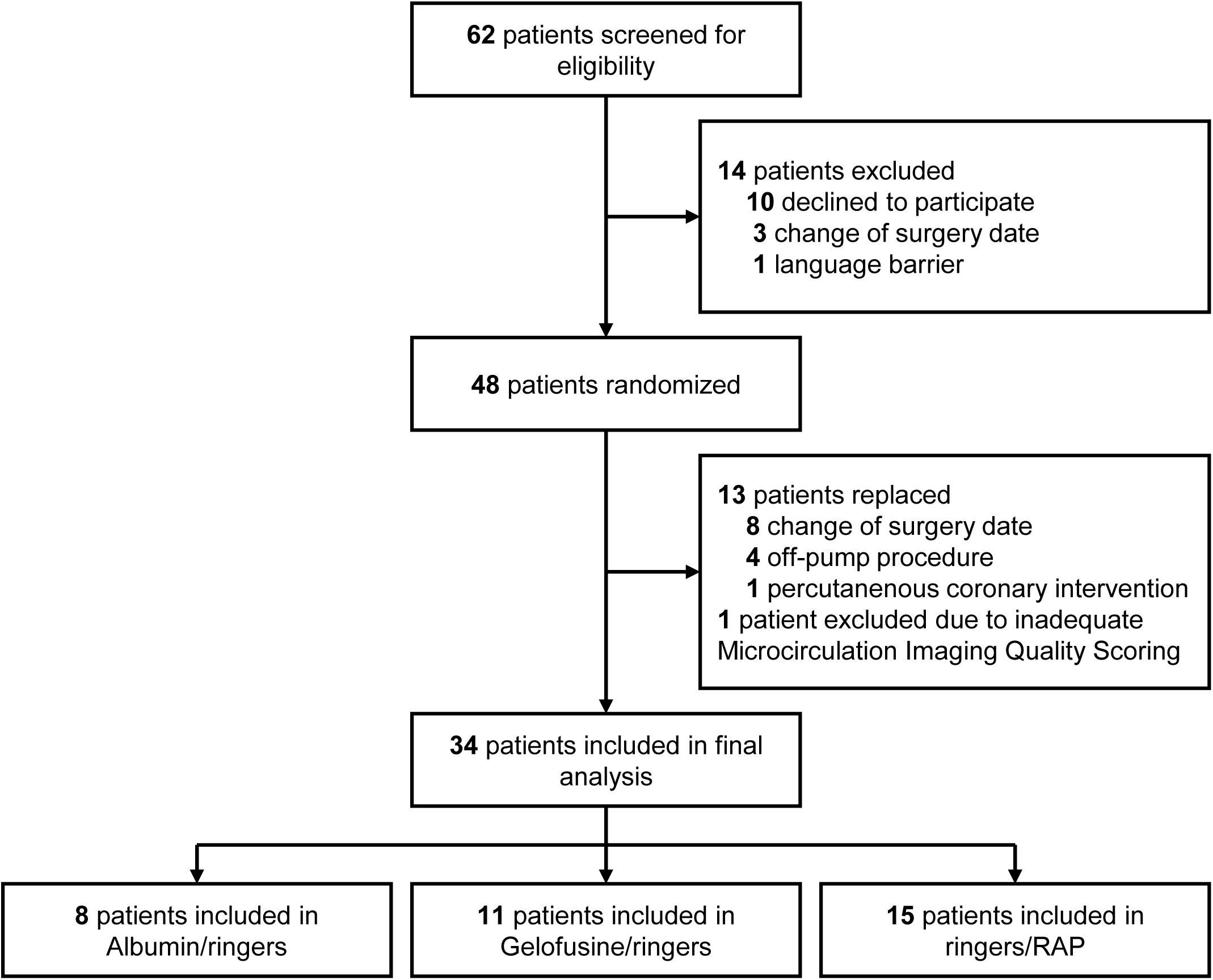

**Table 1.** Patient, and Operative Characteristics.

|  | **Albumin/ ringers**<br>**(n = 8)** | **Gelofusine/**<br>**ringers**<br>**(n = 11)** | **Ringers/RAP**<br>**(n = 15)** |
| --- | --- | --- | --- |
| Sex, Male, n (%) | 6 (75) | 9 (81) | 15 (100) |
| Age, years, median [IQR] | 66 [63-71] | 61 [58-76] | 70 [64-74] |
| BMI, kg/m<sup>2</sup>, median [IQR] | 27.8 [24.5-29.1] | 26.9 [25.7-28.5] | 27.9 [26.7-29.1] |
| BSA, m<sup>2</sup>, median [IQR] | 2.0 [1.9-2.1] | 2.0 [1.8-2.1] | 2.1 [2-2.2] |
| EuroSCORE II, median [IQR] | 0.99 [0.99-0.99] | 0.99 [0.99-1.48] | 0.99 [0.99-1.48] |
| **Comorbidities, n (%)** |  |  |  |
| Current smoker | 2 (25) | 1 (9) | 2 (13) |
| Renal impairment<sup>1</sup> | 1 (13) | 0 | 0 |
| Pulmonary disease | 1 (13) | 0 | 0 |
| Peripheral vascular disease | 1 (13) | 0 | 2 (13) |
| Diabetes Mellitus |  |  |  |
| Type II on insulin | 0 | 3 (27) | 1 (7) |
| Type II without insulin | 1 (12) | 1 (9) | 4 (27) |
| **Preoperative lab, median [IQR]** |  |  |  |
| Hemoglobin, mmol L<sup>-1</sup> | 8.4 [8.2-9.4] | 8.9 [8.2-9.2] | 8.6 [8.4-9.0] |
| Platelet count, 10<sup>9</sup> L<sup>-1</sup> | 272 [198-287] | 197 [178-316] | 215 [201-256] |
| eGFR, mL/min/1.73 m<sup>2</sup> | 81 [75-88] | 89 [72-90] | 84 [72-90] |
| **Preoperative antiplatelet therapy, n (%)** |  |  |  |
| Acetylsalicylic acid | 4 (50) | 10 (91) | 13 (87) |
| Carbasalate calcium | 1 (13) | 1 (9) | 2 (13) |
| DAPT | 2 (25) | 0 | 0 |
| **Operative characteristics** |  |  |  |
| Total surgery time, median [IQR] | 254 [233-306] | 258 [225-282] | 246 [218-288] |
| CPB time, min, median [IQR] | 92 [73-100] | 78 [67-103] | 84 [72-106] |
| AoX time, min, median [IQR] | 68 [45-87] | 56 [47-67] | 54 [49-79] |
| Blood cardioplegia, mmol,<br>median [IQR] | 31 [26-35] | 28 [28-30] | 30 [28-33] |

Means compared with One-way ANOVA. Medians compared with Kruskal-Wallis test.
Frequencies compared with Fisher's Exact test. Abbreviations: ASA, American Society of
Anesthesiologists; AoX, aortic cross clamping; BMI, Body Mass Index; BSA, Body Surface
Area; CPB, Cardiopulmonary Bypass; DAPT, dual antiplatelet therapy; eGFR, estimated
glomerular filtration rate; EuroSCORE II, European System for Cardiac Operative Risk
Evaluation II. <sup>1</sup>Renal impairment is characterized as known chronic kidney disease
preoperatively.

### Perfused Vessel Density

The PVD was comparable between groups at baseline (median 23.8 IQR [19.9-27.1] mm mm^−2^; 20.1 [17.6-25.2] mm mm^−2^; and 21.6 [19.3-24.4] mm mm^−2^ for patients in the albumin/ringers, gelofusine/ringers, and ringers plus RAP group). (Figure 2) PVD decreased after aortic cross clamping in all groups and remained disturbed upon arrival on the ICU. This decrease was most profound in patients receiving albumin/ringers priming (estimated mean difference between baseline and arrival ICU -7.56 95% CI [-11.53 to -3.59] mm mm^−2^), compared to gelofusine/ringers (-4.10 [-7.53 to -0.67] mm mm^−2^), and ringers plus RAP (-3.77 [-6.64 to -0.90] mm mm^−2^), but did not differ between groups.

**Figure 2.**
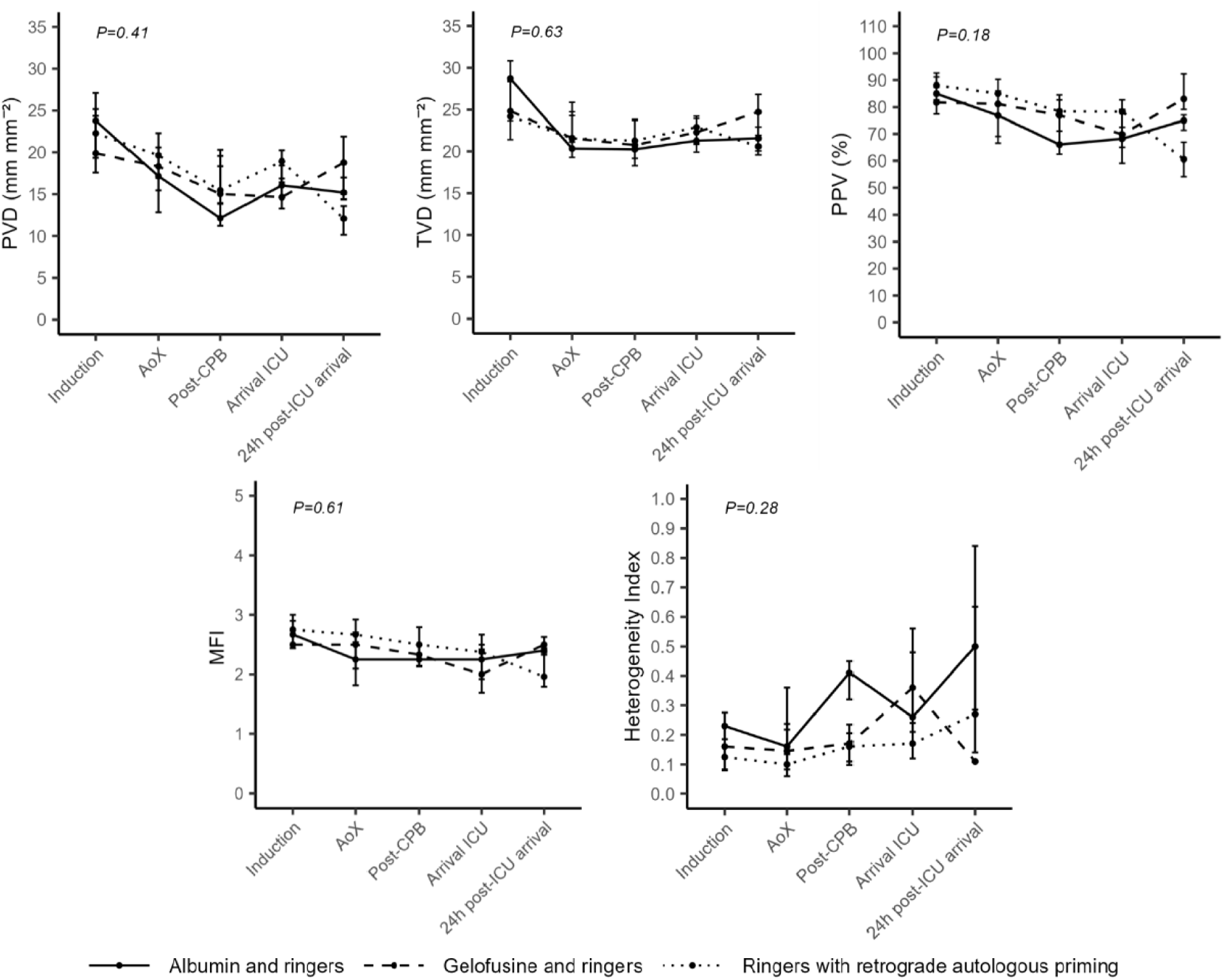
Microcirculatory parameters. Data presented as medians with interquartile ranges. Statistical significance value, P <0.05. Abbreviations: AoX, aortic cross clamping; CPB, cardiopulmonary bypass; ICU, intensive care unit; MFI, microvascular flow index; PVD, perfused vessel density; PPV, proportion of perfused vessels; TVD, total vessel density.

### Microcirculatory perfusion

The TVD decreased after aortic cross clamping in the albumin/ringers and ringers plus RAP groups but not in the gelofusine group. No differences were found between groups over time. For the PPV and MFI, the decrease was significant over time, but not between groups. No differences were found in the heterogeneity index, or the noradrenaline infusion between groups over time COP was comparable after induction of anesthesia in all groups, but decreased after aortic cross clamping in the albumin/ringers and ringers plus RAP group but not in the gelofusine/ringers group. COP partly restored, but did not return to baseline values in all groups. Plasma albumin decreased over time, with differences between groups, and was lowest in patients receiving gelofusine/ringers priming compared to albumin/ringers. Hemoglobin and hematocrit decreased after aortic cross clamping in all groups, and increased upon arrival on the ICU. Hemoglobin levels were highest in patients receiving ringers plus RAP priming. Hemolysis was highest in patients receiving albumin/ringers and differed from patients receiving gelofusine/ringers. (Figure 3 and Table S4)

**Figure 3.**
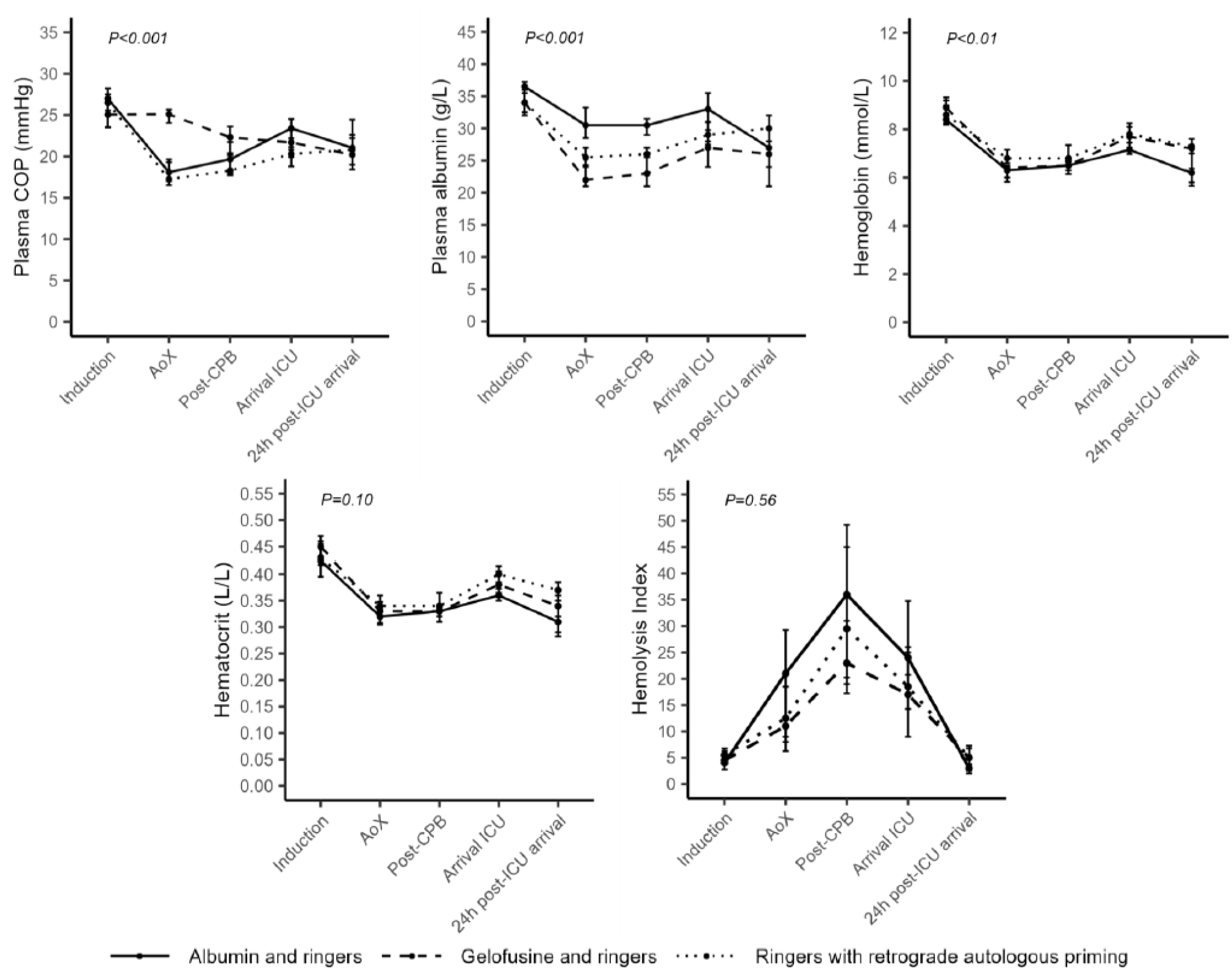
Plasma COP, Albumin, Hemoglobin, Hematocrit and Hemolysis levels. Data presented as median with interquartile range. Statistical significance value, P <0.05. Abbreviations: AoX, aortic cross clamping; CPB, cardiopulmonary bypass; COP, colloid oncotic pressure; ICU, intensive care unit.

Intraoperative fluid requirements and -balances were higher in patients receiving albumin/ringers and ringers plus RAP, compared with gelofusine/ringers. Intraoperative red blood cell transfusions and cell saver volume did not differ between groups. (Table 2)

**Table 2.**
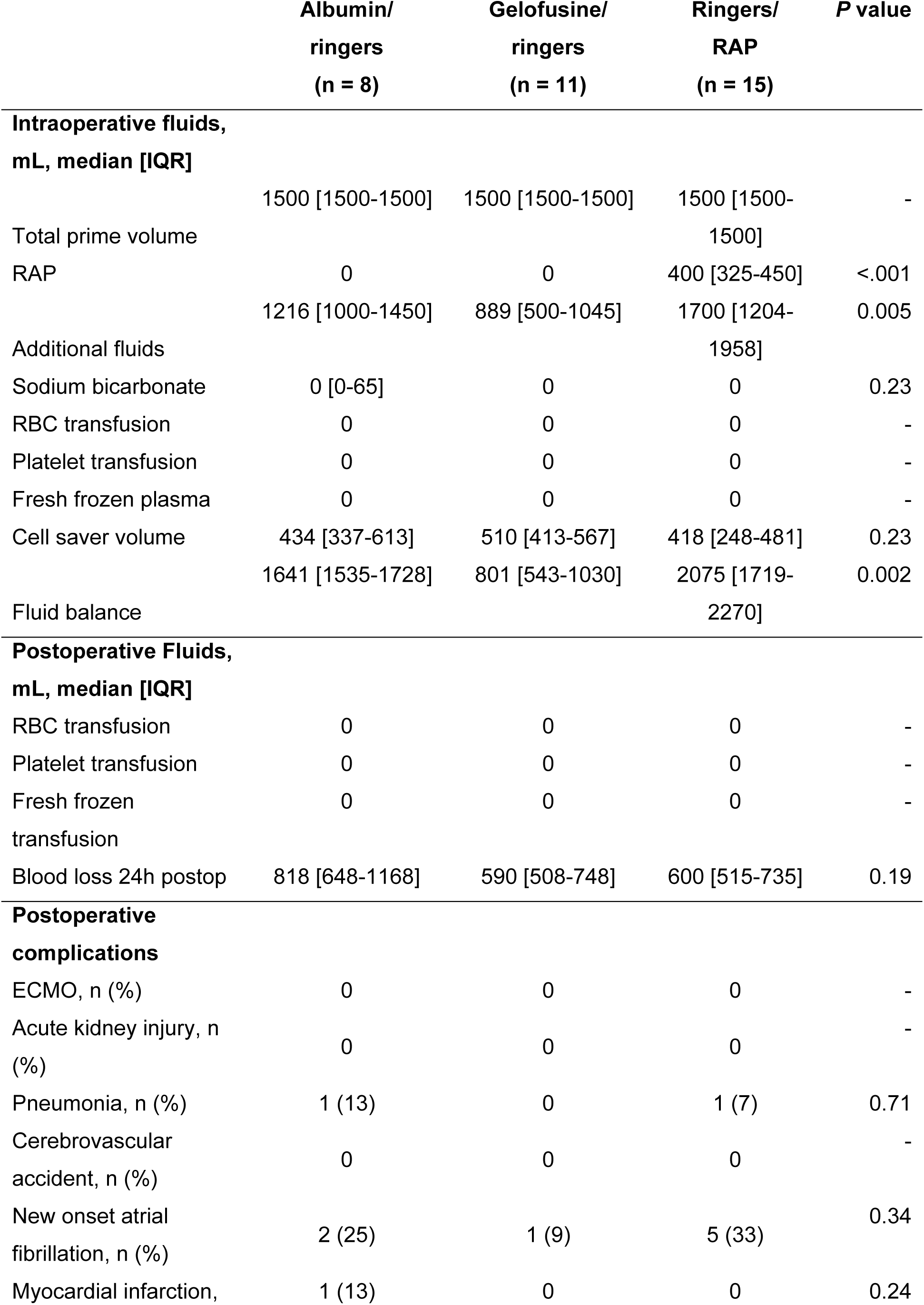
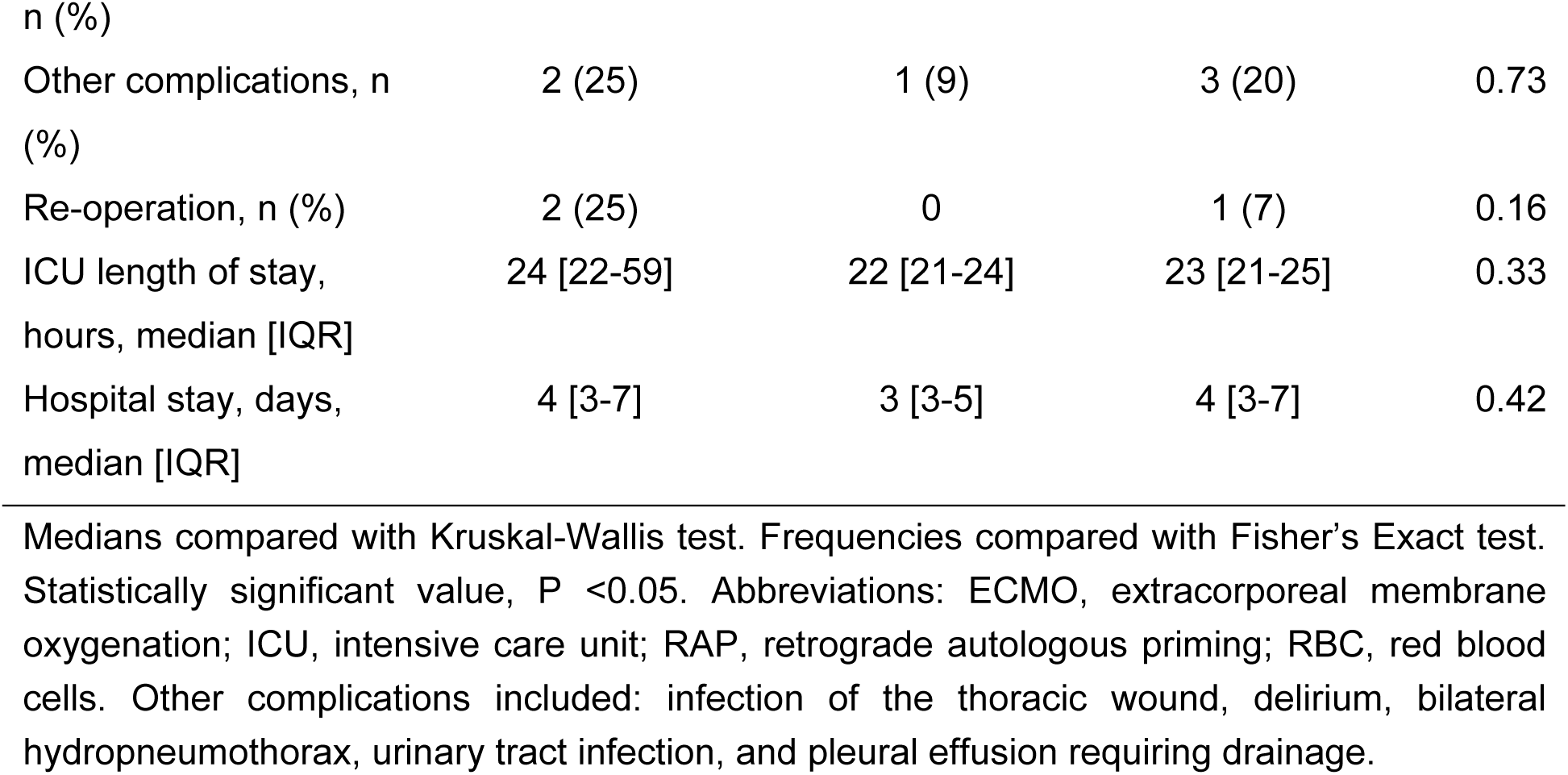
Secondary Outcomes.

Syndecan, heparan sulphate, thrombomodulin, and TNF-α did not differ between groups over time, except for Ang-2, and IL-6. Ang-2 and IL-6 were higher in patients receiving albumin/ringers at 24 hours after ICU admission, compared to those receiving ringers plus RAP. Levels of NGAL were different between groups over time. (Figure 4 and Table S4)

**Figure 4.**
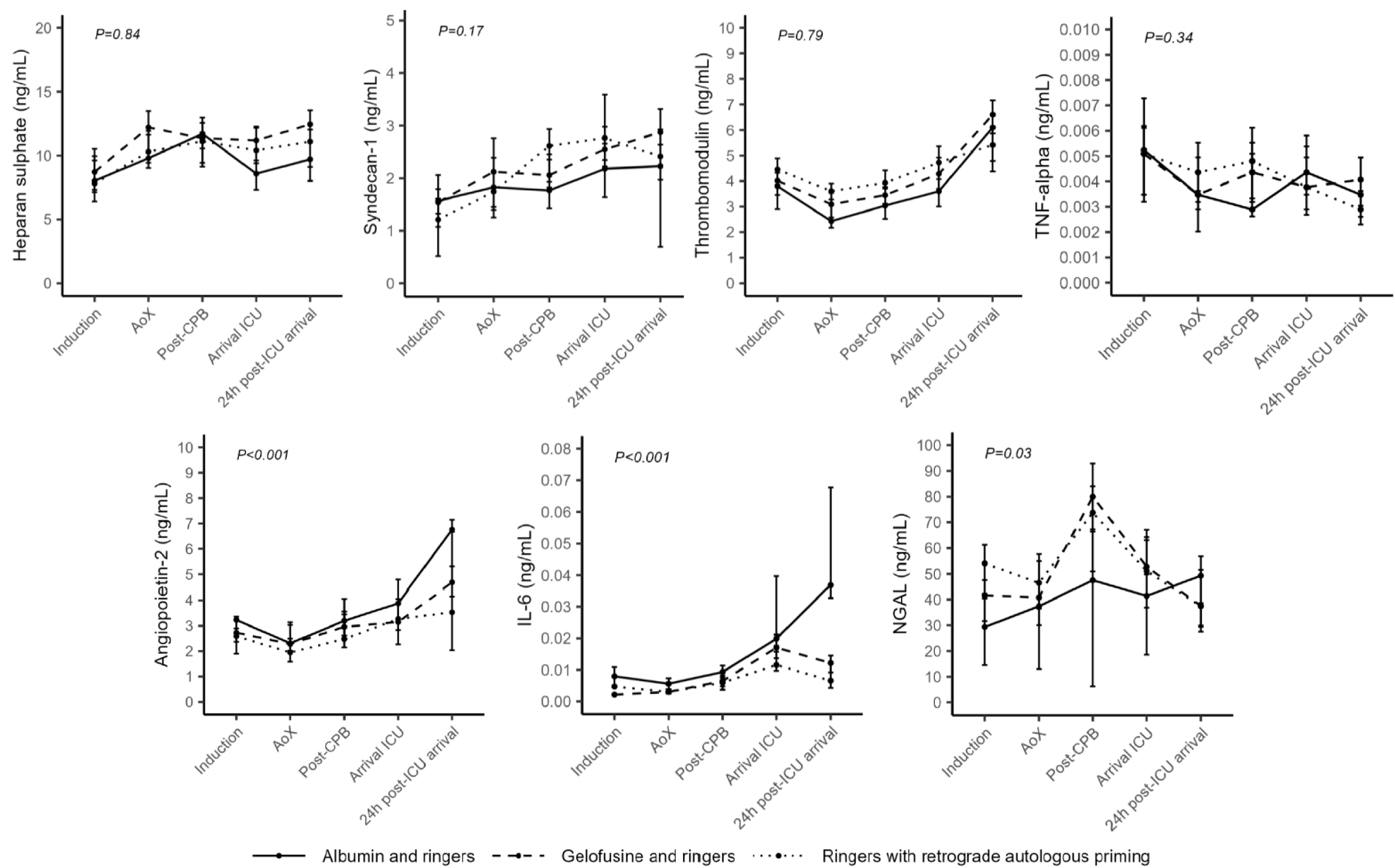
Plasma glycocalyx-, endothelial-, inflammatory-, and renal injury markers. Data presented as median with interquartile range. Statistical significance value, P <0.05. Abbreviations: AoX, aortic cross clamping; CPB, cardiopulmonary bypass; ICU, intensive care unit; IL-6, interleukin-6; NGAL, neutrophil gelatinase-associated lipocalin.

## Discussion

In patients undergoing coronary artery bypass graft surgery with CPB, all the investigated prime fluid strategies - albumin/ringers, gelofusine/ringers or ringers plus RAP – similarly induced perioperative microcirculatory perfusion disturbances. Gelofusine/ringers priming better maintained COP during CPB compared to albumin/ringers and ringers plus RAP, and was paralleled by lower intraoperative fluid balances and less fluid requirements. Additionally, in patients with albumin/ringers, circulating levels of Ang-2 and IL-6 increased 24 hrs after ICU admission.

This study has several strengths. It was designed as a physiological proof-of-principle study to evaluate the effect of CPB priming on microcirculatory perfusion in a translational context, conducted by a team with extensive experience in microcirculatory perfusion assessment. Our group had previously evaluated the effects of CPB priming on microcirculatory perfusion in an animal model, and these findings were incorporated into the study design.^20^ To minimize inter-investigator variability, all investigators performing microcirculatory assessments followed the European guidelines for sublingual microcirculation assessment and received training from experienced researchers, ensuring the reliability of the results.^25^

We observed a decrease in microcirculatory perfusion from the initiation of CPB, without restoration until 24 hours after surgery. This decrease in microcirculatory perfusion during CPB is consistent with previous studies.^7,8,28,29^ However, in our study, none of the investigated prime fluid strategies were able to prevent this decrease in microcirculatory perfusion following CPB. Previous studies have suggested that colloids, such as albumin or gelofusine, may better preserve microcirculatory perfusion than crystalloids in septic patients and animals in hemorrhagic shock.^30–33^ However, our study failed to replicate these findings in the context of CPB. In contrast to our results, other studies reported reduced organ injury with colloid-based CPB priming. A prospective study reported tthat albumin as additive in CPB priming reduced pulmonary edema in rats on CPB compared with hydroxyethyl starch (HES).^20^ Additionally, a randomized controlled trial showed reduced pleural effusion in patients following cardiac surgery receiving albumin-based CPB priming compared with crystalloids alone.^34^ While, colloid-based CPB prime fluid strategies show potential benefits in terms of organ dysfunction, the evidence remains conflicting. A retrospective study reported a comparable incidence in AKI following cardiac surgery in patients receiving a HES- or and gelatin based CPB priming.^35^ Moreover, the use of albumin as CPB priming did not reduce the incidence of AKI or major adverse events compared to crystalloids following cardiac surgery.^19,36^

Although, we hypothesised albumin to protect microcirculatory integrity, based on previous results, patients receiving albumin/ringers exhibited more endothelial damage and inflammation following cardiac surgery compared to those receiving gelofusine/ringers, and ringers plus RAP.^20,37,38^ However, since this study was not powered to detect differences in endothelial-, glycocalyx-, or inflammatory markers, these findings should be considered exploratory. While microcirculatory dysfunction following sepsis and CPB shares several key mechanisms, (e.g., systemic inflammation, glycocalyx degradation, and increased vascular permeability), a potential difference could lie in the hemodilution at CPB initiation.^15,29,39,40^ While moderate hemodilution is often well tolerated though compensatory mechanisms, severe hemodilution of CPB priming may become pathophysiological, comprosmising microcirculatory function.^10,41^

In this study, COP was better preserved after aortic cross-clamping in patients receiving gelofusine/ringers compared to those receiving albumin/ringers or ringers plus RAP. This was accompanied by lower intraoperative fluid requirements and fluid balances. Intraoperative fluid requirements, fluid balances, and hemoglobin levels were highest in patients receiving ringers plus RAP, suggesting greater interstitial fluid accumulation in this group. These findings might indicate potential differences in microcirculatory behavior between the groups, though microcirculatory perfusion was equally disturbed. The impact of hemodilution may play a greater role than the composition of CPB priming in affecting microcirculatory perfusion. A previous showed that using a mini-CPB system better preserved microcirculatory perfusion compared to conventional CPB, suggesting that prime fluid composition might be overshadowed by the impact of hemodilution at CPB initiation, a key factor attributing to microcirculatory impairment.^10,42^

Our findings suggest that gelofusine/ringers as a CPB priming strategy may be a viable option for preserving COP and preventing interstitial fluid accumulation, without negatively impacting microcirculatory integrity. Compared to albumin/ringers and ringers plus RAP, gelofusine/ringers may offer an advantage in preventing interstitial fluid accumulation, potentially contributing to the preservation of organ function following cardiac surgery.

This study has several limitations. First, it was a single-center trial conducted in a tertiary university hospital, reducing the generalizability of the findings. Second, RAP was performed only in the prime composition consisting solely of ringers, as our goal was to assess the impact of the prime solution itself—rather than volume—on microcirculatory perfusion. We did not evaluate a priming strategy consisting exclusively of ringers without RAP, as the combination of ringers plus RAP (rather than ringers alone) is the common priming strategy used in the Netherlands. Patients receiving ringers plus RAP experienced less initial hemodilution compared to other groups, potentially influencing our results. Nevertheless, the RAP volume was displaced during CPB in all patients.Third, the use of mannitol was common practice in our hospital. Fourth, there was a high rate of missing data 24 hrs post-surgery, primarily due to patients declining further microcirculatory assessments. While data at other time points were sufficient, the missing data may have contributed to increased variability. Fifth, variability in microcirculatory perfusion within groups suggests the sample size may have been insufficient. Sixth, the study population was limited to patients undergoing coronary artery bypass graft surgery, reducing the generalizability of the results for all cardiac surgery patients.

## Conclusion

In patients undergoing coronary artery bypass graft surgery with cardiopulmonary bypass, all of the studied prime fluid strategies - albumin/ringers, gelofusine/ringers, or ringers plus RAP – similarly induced microcirculatory perfusion disturbances. Although, gelofusine/ringers better maintained colloid oncotic pressure and resulted in lower intraoperative fluid requirements and fluid balances, no differences in microcirculatory perfusion were observed between groups. Albumin priming resulted in increased inflammatory markers at 24 hrs after surgery. These findings highlight the complexity of preserving microcirculatory integrity during cardiac surgery.

## Data Availability

The data used during the current study are available from the corresponding author on reasonable request.

## Non-standard Abbreviations and Acronyms

Ang-2: Angiopoietin-2
BSA: Body Surface Area
CABG: Coronary Artery Bypass Graft
CPB: CardioPulmonary Bypass
COP: Colloid Oncotic Pressure
HES: Hydroxyethyl starch
ICU: Intensive Care Unit
IL-6: Interleukin-6
MFI: Microvascular Flow Index
NGAL: Neutrophil Gelatinase-Associated Lipocalin
PPV: Proportion of Perfused Vessels
PVD: Perfused Vessel Density
RAP: Retrograde Autologous Priming
SDF: Sidestream Darkfield Imaging
TNF- α: Tumor Necrosis Factor Alpha
TVD: Total Vessel Density

## Acknowledgements

Not applicable.

